# AMYLASE DEPLETION SIGNIFICANTLY IMPROVES THE MALDI-TOF PROFILE OF SALIVA – APPLICATION TO COVID-19 DIAGNOSTICS

**DOI:** 10.1101/2023.04.11.23288361

**Authors:** Zane LaCasse, Prajkta Chivte, Kari Kress, Venkata Devesh R. Seethi, Joshua Bland, Hamed Alhoori, Shrihari S. Kadkol, Elizabeth R. Gaillard

**Affiliations:** Departments of Chemistry and Biochemistry, Northern Illinois University, DeKalb, IL 60115; Departments of Computer Science, Northern Illinois University, DeKalb, IL 60115; Thermo Fisher Scientific, Rockford, IL 61101; Department of Pathology, University of Illinois at Chicago, Chicago, IL 60612

**Author notes:** co-first authors (Zane LaCasse, Prajkta Chivte).

**Keywords:** Saliva, MALDI-ToF, Amylase, high-abundance, low-abundance, COVID-19

## Abstract

Human saliva contains a plethora of proteins whose presence and concentration can be monitored for diagnosis and progression of disease. Saliva has been extensively probed for the diagnosis of several systemic and infectious diseases because of the ease with which it can be collected. However, amylase, the most abundant protein found in saliva can obscure the detection of low-abundance proteins by MALDI-ToF MS (matrix-assisted laser desorption/ionization-time of flight mass spectrometry) and diminish the diagnostic utility of this specimen type. In the present study, we used a device to deplete salivary amylase from water-gargle samples through affinity adsorption. After depletion, profiling of the saliva proteome was performed by MALDI-ToF MS on gargle samples from subjects whose COVID-19 (coronavirus disease 2019) status was confirmed by NP (nasopharyngeal) swab RT-qPCR (reverse transcription polymerase chain reaction). Amylase depletion led to the enhancement of signal intensities of various peaks as well as the detection of previously unobserved peaks in the MALDI-ToF spectra. The overall specificity and sensitivity after amylase depletion was 100% and 85.17% respectively for detecting COVID-19. Our simple, rapid and inexpensive technique to deplete salivary amylase can be used to unmask spectral diversity in saliva by MALDI-ToF MS, reveal low-abundant proteins and aid in the establishment of novel biomarkers for diseases.

## Introduction

Saliva is a complex biological fluid consisting of a mixture of major and minor salivary gland secretions, nasal and bronchial secretions, plasma filtrates, host cells and bacteria. The fluid is slightly acidic and contains various organic (proteins, hormones, nucleic acids, fatty acids) and inorganic (Na^+^, K^+^, Cl^−^, Ca_2_ ^+^, HCO _3_^−^, H _2_ PO _4_^−^, I^−^, Mg _2_^+^ and NH _4_^+^) components.[1],[2],[3],[4]

Described as a “mirror of the body,” saliva reflects the physiological and pathological states of the body. Saliva has been used for screening[5],[6] diagnosis [7],[8] prognosis and monitoring[9],[10] of numerous human diseases. Collecting saliva is fast, non-invasive and can be done by the subjects themselves. Saliva does not need any transport medium and requires minimal processing before testing compared to other specimen types.[11],[12] Major advancements in the field of salivary diagnostics include RNA-sequencing, point-of-care technologies, liquid biopsies and protein profiling.[13]

Proteomic analysis of saliva has led to the discovery of ∼3000 proteins, including mucins, proline-rich proteins, histatins, statherins, cystatins, interleukins, amylase, albumin and immunoglobulins (among others).[3],[14] A number of these proteins are currently being pursued as biomarkers for oral diseases[15],[16], various cancers[17],[18] Srojen’s syndrome[19], and even autism[20]. Saliva has been used to detect viruses such as hepatitis B virus[21], human immunodeficiency virus[22], dengue[23] and, more recently, severe acute respiratory syndrome coronavirus type-2 (SARS-CoV-2)[1],[4],[24]. Several studies have demonstrated higher efficiencies of SARS-CoV-2 detection in saliva compared to nasopharyngeal swabs, owing to the fact that the receptor used for SARS-COV-2 viral entry, angiotensin-converting enzyme-2 (ACE-2), is abundantly expressed in the oral epithelial cells.[25],[26],[27] Some studies have shown that SARS-CoV-2 can be detected 1-5 days earlier in saliva samples when compared to nasopharyngeal swabs.[28]

A major hurdle in proteomic analyses of saliva, however, is the presence of high-abundance proteins that can mask the presence of target proteins expressed at a lower level. Salivary α-amylases, albumin and immunoglobulins alone make up to 75% of the total saliva proteome. Consequently, it is crucial to deplete these high-abundance proteins (or conversely, to enrich low-abundance proteins) to improve detection sensitivity.[1],[3],[29] For instance, non-analyte, high-abundance proteins can cause significant ion suppression for low-abundant biomarkers in mass spectrometric techniques. Particularly, in a complex biological matrix, the ionization efficiencies are highly influenced by the abundance, molecular weight and chemistry of the proteins.[30]

Salivary α-amylase is the most abundant of these proteins and is an enzyme that catalyzes the hydrolysis of 1,4-glucosidic linkages in starch and other polysaccharides. It exists in two proteoforms: non-glycosylated (∼56 kDa) and glycosylated (∼59 kDa).[31],[32],[33] Several techniques are currently utilized for α-amylase depletion including a syringe-based potato starch device[34], gel filtration[35], affinity chromatography by lectin ConA[31], co-precipitation and paper-based chips[36]. Deutsch et al. designed and patented an amylase depleting device to separate salivary α-amylase from the whole saliva by affinity adsorption to potato starch.[34] The authors demonstrated a six-fold amylase reduction as compared to the total amylase in the saliva, and 97% reduced amylase activity after using this device. [34] Xiao et al. and Crosara et al. validated the functionality and practicality of this device via SDS-PAGE and western blotting. In the study, several identified proteins, which could serve as potential biomarkers, were unequivocally detected after amylase removal including desmoplakin, short palate lung and nasal epithelium carcinoma-associated protein 2, mucin-7 and several immunoglobulins’ isoforms.[37], [2]

In our previous study, we demonstrated the utility of matrix-assisted laser desorption-ionization time of flight mass spectrometry (MALDI-ToF MS) to distinguish COVID-19 (coronavirus disease 2019) positive and negative individuals.

[38] One hurdle in the analysis of the protein profiles was the dominant signal of salivary amylase resulting in ion suppression of other proteins. We hypothesize that depletion of salivary α-amylase from gargle samples before MALDI-ToF analysis would unmask and improve the detection of low-abundance proteins that are otherwise difficult to visualize. The simple removal of amylase coupled with the sensitivity of MALDI-ToF could open avenues for the discovery of new protein biomarkers in saliva-based diagnostics. Taking this into account, we combined methods to remove amylase by affinity adsorption with potato starch and salivary protein profiling using MALDI-ToF MS. We also optimized sample preparation involving the viral disruption buffer. In our previous study, we used LBSD-X buffer by MAPSciences (Bedford, UK) as a viral disruption buffer for gargle samples to test for COVID-19. However, LBSD-X buffer is not readily available, and the exact composition of this buffer remains unknown. Hence, the viral disruption buffer reported by Dollman et al. was used in this study. The buffer has already been used in methods for the detection of SARS-CoV-2 and influenza viruses such as H1N1 and H3N2.[39],[40],[41] Overall, we aimed to develop a simple, rapid and inexpensive technique to deplete salivary amylase and to detect COVID-19 by performing MALDI-ToF protein profiling.

## Materials and Method

### Ethical and Biosafety statement

The study was approved by the Institutional Review Board and Institutional Biosafety Committee of Northern Illinois University (approved on August 12, 2020, and revised on July 12, 2021, and August 11, 2022). Informed consent was obtained from the volunteers who participated in the study. The collected information was strictly limited to demographics, COVID-19 vaccination status, symptoms, and RT-qPCR results. Sample handling and processing were conducted under a Class II Biosafety Cabinet. All methods were performed in accordance with the relevant guidelines and regulations.

### Sample collection

Samples were collected at drive-thru testing sites conducted by the Illinois Department of Public Health (IDPH) at Aurora and Rockford, Illinois between June 2021 and July 2022. At these sites, an NP swab was collected and analyzed by RT-qPCR to detect SARS-CoV-2 at an IDPH laboratory. RT-qPCR results were reported as Detected, Not detected, or Inconclusive. Individuals who consented to participate in the research study were asked to provide a water gargle sample at the same time as the NP swab sample collection. Subjects were asked to gargle 10 mL of bottled spring water for 30 s, which was then collected in a 50 mL conical centrifuge tube. The samples were stored at − 20 °C until processing and analysis. A total of 107 gargle samples were collected to ensure that a substantial pool of COVID-19 positive samples was included. For the current analysis, gargles from 14 COVID-19 positive and negative subjects were analyzed retrospectively by MALDI-TOF MS after amylase depletion.

### Preparation of samples and controls

#### i. Gargle samples

The gargle samples were thawed and transferred to a 30 mL disposable polypropylene beaker (Fisher Scientific, Waltham, MA). Samples were filtered through a 0.45 µm polyethersulfone membrane filter (Celltreat Scientific Products, Pepperell, MA), divided equally and transferred into two 50 mL tubes (approximately 5 mL of gargle samples in each tube).

### Amylase removal using potato starch

The filtered gargle sample from one tube was hand-pressed through an amylase depleting device, with modifications to a previously described apparatus.[34] The device comprised a 10 mL plastic syringe (Thermo scientific, Rockwood, TN) with a 0.45 µm syringe filter fixed at the tip (Figure 1).

**Figure 1:**
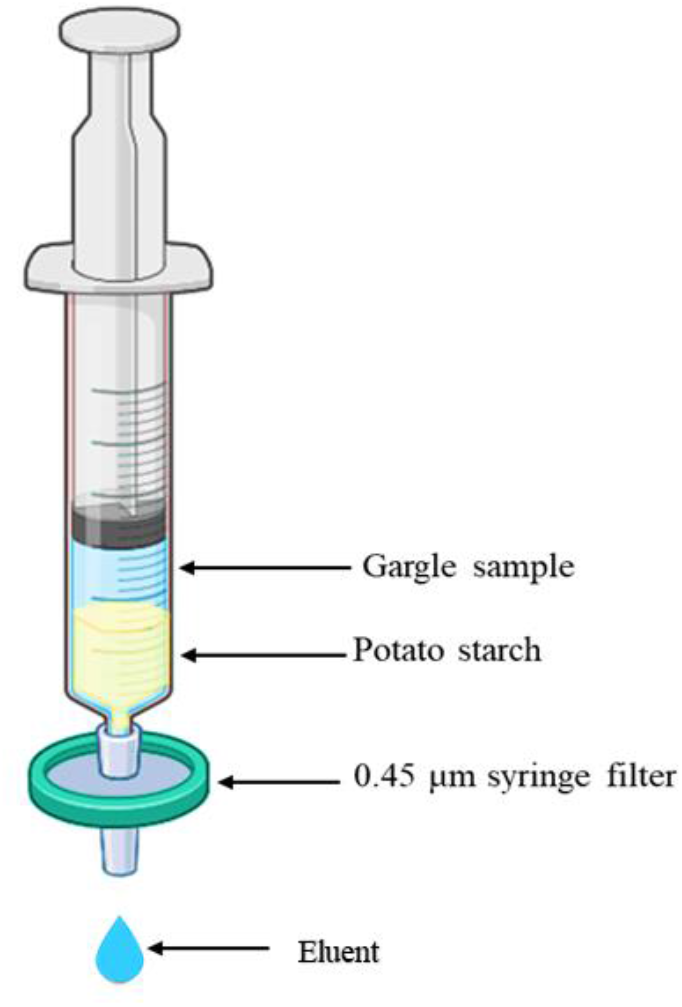
An illustration of salivary amylase depleting device loaded with a gargle sample (Created with BioRender.com)

The syringe was filled with a slurry of 2 g of potato starch (Sigma-Aldrich, St. Louis, MO) in 20 mL of LC-MS grade H_2_O (OmniSolv, Sigma-Aldrich, St. Louis, MO). The slurry was hand-pressed through the setup to moisten the substrate and remove any water-soluble residues. To optimize the amount of water required to wash off all water-soluble starch residues, we passed 5 mL of LC-MS H_2_O (30 mL total) through the amylase depleting device and collected the respective filtrates. An iodine reagent (0.15 M I_2_ + 0.3 M KI;10 μL) was added to 1 mL of each filtrate and the absorbance was measured using a UV-Vis spectrometer (UV-2600, Shimadzu). Thereafter, 5 mL of the filtered gargle sample was hand-pressed and filtered through the slurry filled syringe column. The resultant 5 mL of eluent was collected in a new 50 mL tube (Figure 1). Next, 5 mL of chilled acetone (Sigma-Aldrich, St. Louis, MO) was added to precipitate proteins. Acetone was added directly to the other aliquot that did not go through amylase depletion. The samples were centrifuged in a Beckman Coulter Avanti J-E series centrifuge with a JA-20 rotor at 16,000 × g for 30 min at 4°C. The supernatant was discarded, the rim of the tube patted dry, and the pellet resuspended in 50 μL of a viral disruption buffer reported by Dollman et al. consisting of 50 mM ammonium bicarbonate (Sigma-Aldrich, St. Louis, MO), 10% acetonitrile (Oakwood Chemical, Estill, SC), 50 mM TCEP (Sigma-Aldrich, St. Louis, MO), and 5 mM octyl ß-D-glucopyranoside (Sigma-Aldrich, St. Louis, MO) at pH 7.5. [39] The reconstituted pellets were vortexed for 30 s and incubated at room temperature for 15 min.

#### ii. Standards and controls

A negative control, consisting of pooled human saliva (pre-COVID-19) collected before November 2019 (Lee Biosolutions, Maryland Heights, MO), was prepared by spiking 500 µL thawed stock into 10 mL of water. The control was filtered and processed as a gargle sample and subjected to amylase depletion as described before. Two hundred pmol of human serum IgA, 200 pmol of human serum albumin and 2000 pmol of human amylase (Sigma-Aldrich, St. Louis, MO) in LC-MS grade H_2_O were also passed through the amylase depleting device. The protein standards and their respective eluents were then lyophilized overnight and reconstituted in 10 µL of viral disruption buffer. A final concentration of 20 pmol, 20 pmol and 200 pmol of IgA, HSA and amylase were spotted for MALDI-ToF analysis, respectively.

### Sample Spotting

The sandwich method (1 µL matrix-1 µL sample-1 µL matrix) was employed for spotting the samples on the MALDI-MS target plate (Shimadzu, Kyoto, Japan). Sinapinic acid (Sigma-Aldrich, St. Louis, MO) was used as the matrix with a concentration of 20 mg/mL in a 1:1 LC-MS grade H_2_O: Acetonitrile solution containing 0.1% trifluoroacetic acid (Sigma-Aldrich, St. Louis, MO). The matrix was freshly prepared every seven days and stored at 4°C between analyses.

### Instrument parameters

Shimadzu AXIMA Performance MALDI-ToF MS (Shimadzu Kratos Analytical, Manchester, UK) was used for MS acquisition. It was equipped with a nitrogen laser set at 337.1 nm with a pulse width of 3 ns and a maximum repetition rate of 60 Hz. The AXIMA Performance mass spectrometer was operated with Shimadzu Biotech Launchpad Software (version 2.9.4) and was run in positive-ion linear detection mode. The laser power and repetition rate were set to 100 µJ/pulse and 50 Hz, respectively. The spectra were acquired in the range of 2,000–200,00 m/z by summing 5000 spectra (250 profiles by 20 shots). Pulse extraction was set to 50,000 m/z and the ion gate was set to blank values below 1500 m/z. Post-acquisition baseline subtraction and smoothing (Gaussian) were performed using Shimadzu Biotech Launchpad software.

### Instrument calibration

The instrument was calibrated daily using the (M+H)^+^ and (M+2H)^2+^ peaks of ProteoMass Apomyoglobin MALDI-MS Standard (Sigma-Aldrich, St. Louis, MS), prepared at 100 pmol/µL in LC-MS grade H_2_O. The signal intensities of the calibrant were recorded throughout the entire analysis to track the inter-day instrument performance. Calibration was accepted if the mass deviation was less than 500 mDa.

### Data analysis

Shimadzu Biotech Launchpad was used to export a text file for each gargle sample that was passed through the amylase depleting device, which included the spectrum of mass-to-charge values ranging from 2,000 m/z to 200,000 m/z alongside the respective ion count intensities. The area under the curve (AUC) was calculated as described in our previous study. In brief, the previously established biomarker ranges (11,140–11,160 m/z, 23,550–23,800 m/z, 27,900–29,400 m/z, 55,500– 59,000 m/z, 66,400–68,100 m/z, 78,600–80,500 m/z, 111,500–115,500 m/z) were presumably indicative of host immune proteins or viral proteins. The intermittent values of these ranges indicated the presence of protein masses; hence ion counts in each subrange were coalesced together through the integration of points to calculate the AUC which produced features for each data sample. AUC values were computed by leveraging the composite Simpson’s rule for each spectral range.[38] The peak at 11,140–11,160 m/z was used as an internal control to verify whether a given gargle sample had been successfully collected and processed. The areas of 11,140–11,160 m/z and 78,600–80,500 m/z (the approximate theoretical m/z of the S1 fragment of the SARS-CoV-2 spike protein) peaks were calculated for all the samples that were passed through the amylase depleting device. Receiver Operating Characteristic (ROC) curve analysis was done to determine the sensitivities and specificities (Medcalc software, Belgium). The true positive rate (sensitivity) on the Y-axis was plotted against the false positive rate (100-specificity) on the X-axis for each of the observed AUCs for the peaks at 11,140–11,160 m/z and 78,600– 80,500 m/z.

### SDS-PAGE analysis

To determine the efficiency of amylase depletion, the remaining reconstituted pellets were run on 8% Bis-Tris Plus acrylamide gels (Invitrogen, Waltham, MA) for 35 min at 150 V with MES SDS running buffer (Novex, Waltham, MA). Gargle samples (20 µL) before and after passing through the amylase depleting device and protein standards of amylase (14 µg), HSA (2.5 µg) and IgA (3.5 µg) were analyzed on the gels under reducing conditions. The gels were stained using a 0.1% w/v PhastGel Blue R (Amersham Biosciences, Uppsala Sweden) solution for 20 min, followed by de-staining with 30% v/v methanol (ASC grade) and 10% v/v acetic acid (ACS grade) in Milli-Q water for 45 min. Protein bands were visualized to determine the efficiency of amylase depletion.

## Results

### The starch bed selectively binds amylase

Before testing the amylase depleting device with gargle samples from subjects, we determined whether the procedure affected the detection of other proteins found in saliva. Individual protein standards that exist in high-abundance in saliva, including salivary amylase, HSA and IgA were tested. When standard amylase was passed through the device, the collected eluent showed a negligible signal at the theoretical m/z value of the parent peak confirming efficient removal of amylase (Figure 2). Moreover, as anticipated, when HSA and IgA were passed through this device, minimal to no binding/interaction was observed between the starch bed and these two proteins (Figures 3 and 4). The spectra from these experiments confirmed that the starch bed selectively binds amylase and results in negligible sample loss of other proteins. Next, we passed pre-COVID-19 saliva, the negative control, through this device. The complete protein profile of the negative control before and after passing through the amylase depleting device has observable differences (Figure 5). For instance, the intensity of the peak at 56,216 m/z (approximately the theoretical molecular weight of salivary amylase) was considerably reduced after the sample was passed through the device whereas the peak around 66,000 m/z was enhanced. A low intensity peak between 56,000-58,000 m/z was maintained in the eluent. Comparing Figures 2 and 4, this signal coincides with the heavy chain of salivary IgA, which would not be retained on the starch bed.

**Figure 2:**
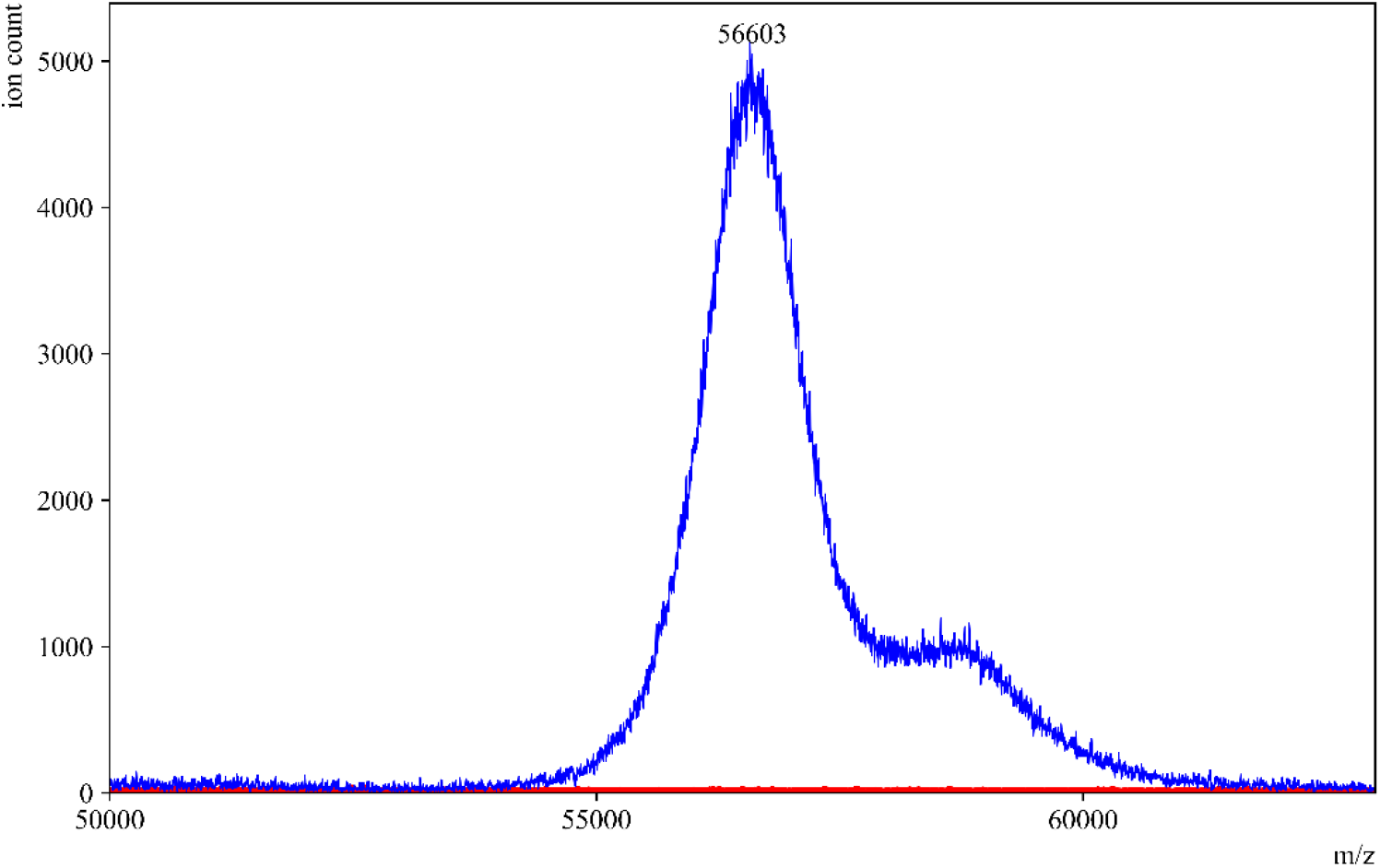
Standard amylase before and after passing through amylase depleting device. MALDI-ToF mass spectra of standard amylase in (—blue) and an overlay of the same sample after passing through the amylase depleting device (—red). No peak was observed at 56,000 m/z after passing through the device, confirming the binding between the starch bed and salivary amylase.

**Figure 3:**
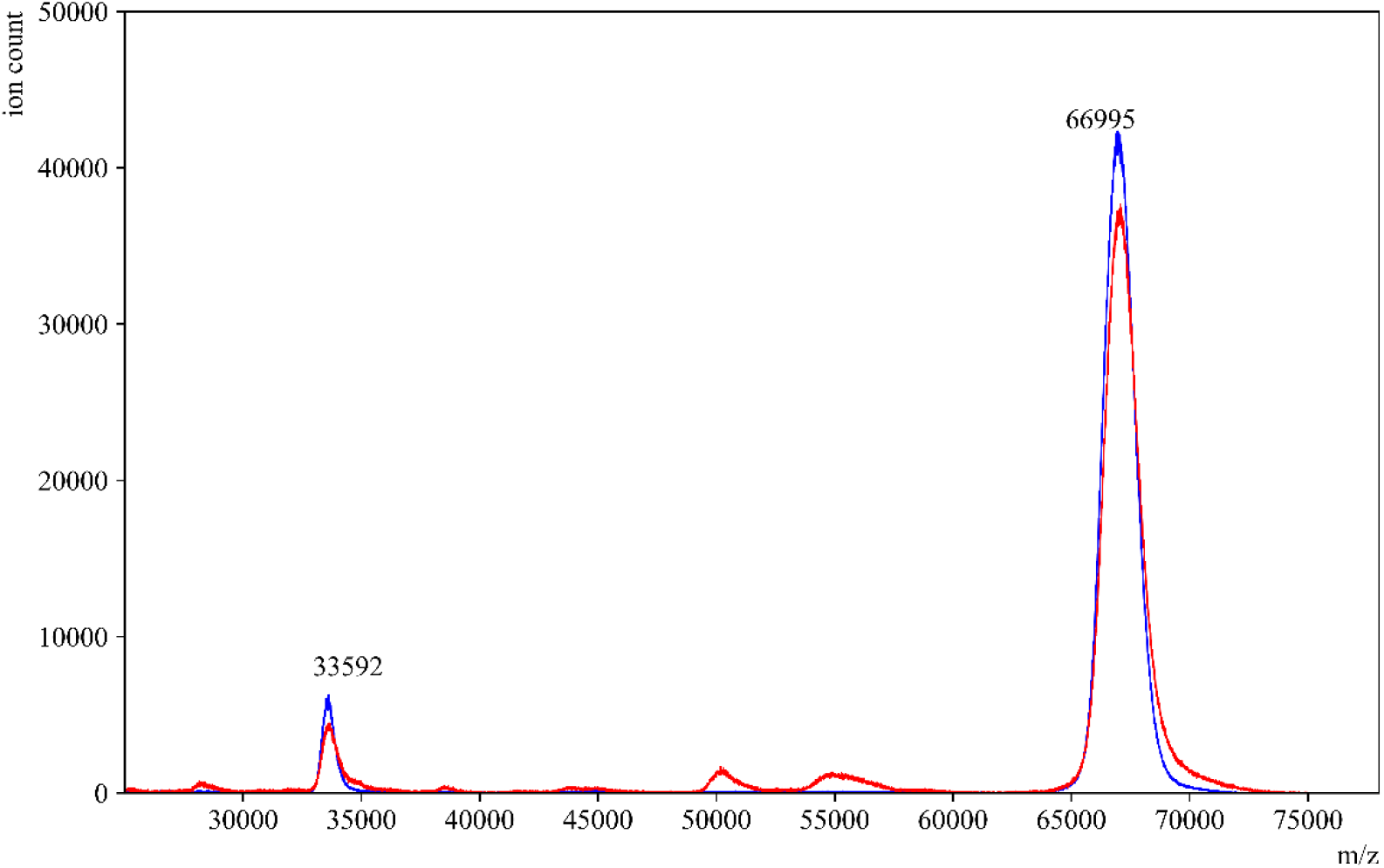
Standard HSA before and after passing through amylase depleting device. MALDI-ToF mass spectra of standard HAS in (—blue) and an overlay of the same sample after passing through the amylase depleting device (—red). The peaks at 66,995 m/z and 33,592 m/z which correspond to the single- and double-charged peaks of HSA, respectively, were observed with similar signal intensities before and after using the amylase depleting device.

**Figure 4:**
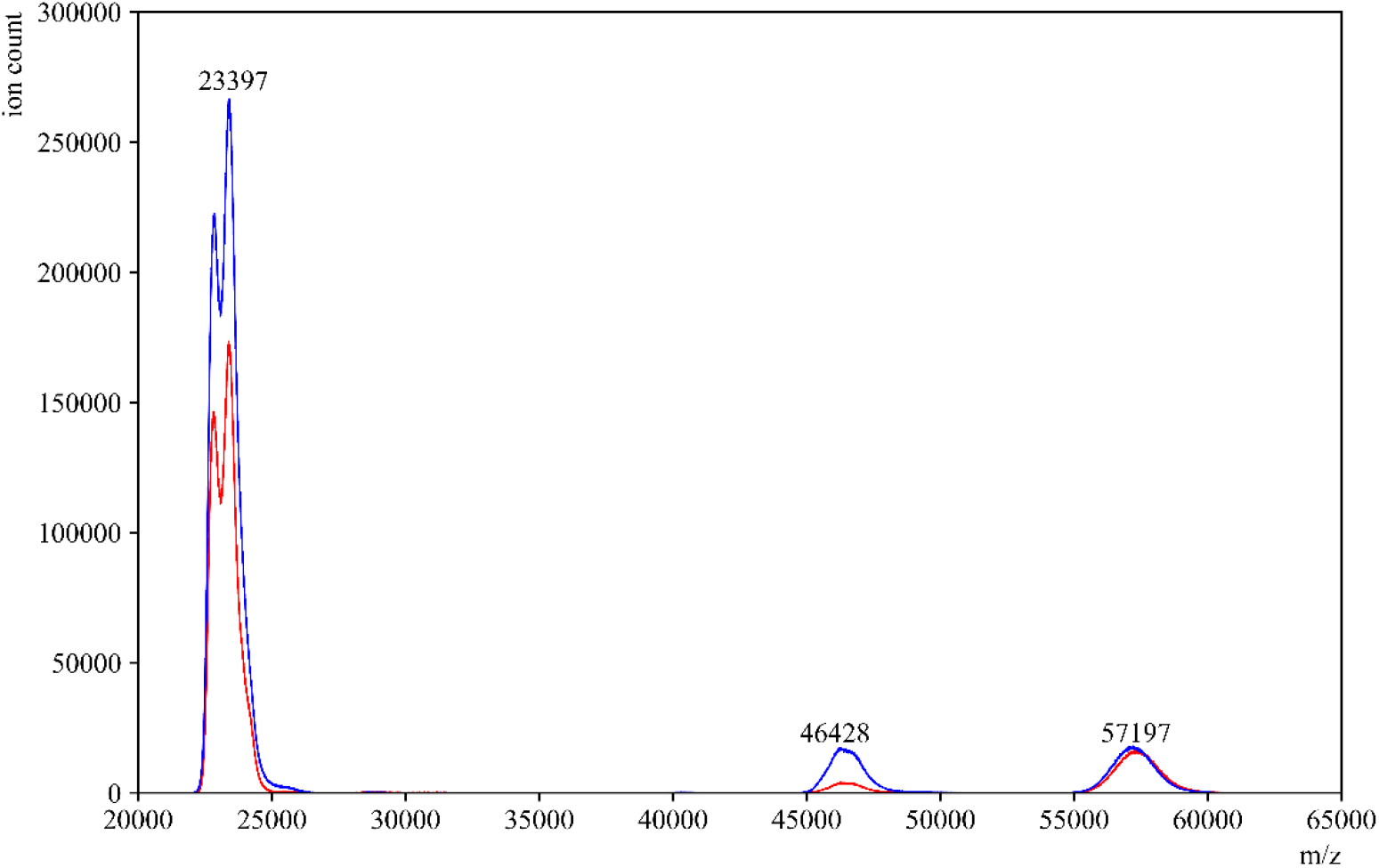
Standard IgA before and after passing through amylase depleting device. MALDI-ToF mass spectra of a standard human serum IgA in (—blue) and an overlay of the same sample after passing through the amylase depleting device (—red). The peaks at 23,397 and 57,197 m/z which correspond to the light chain and heavy chain peaks of IgA respectively, can be observed with similar signal intensities before and after using the amylase depleting device. The peak at 46,428 m/z is the dimer of the IgA light chains.

**Figure 5:**
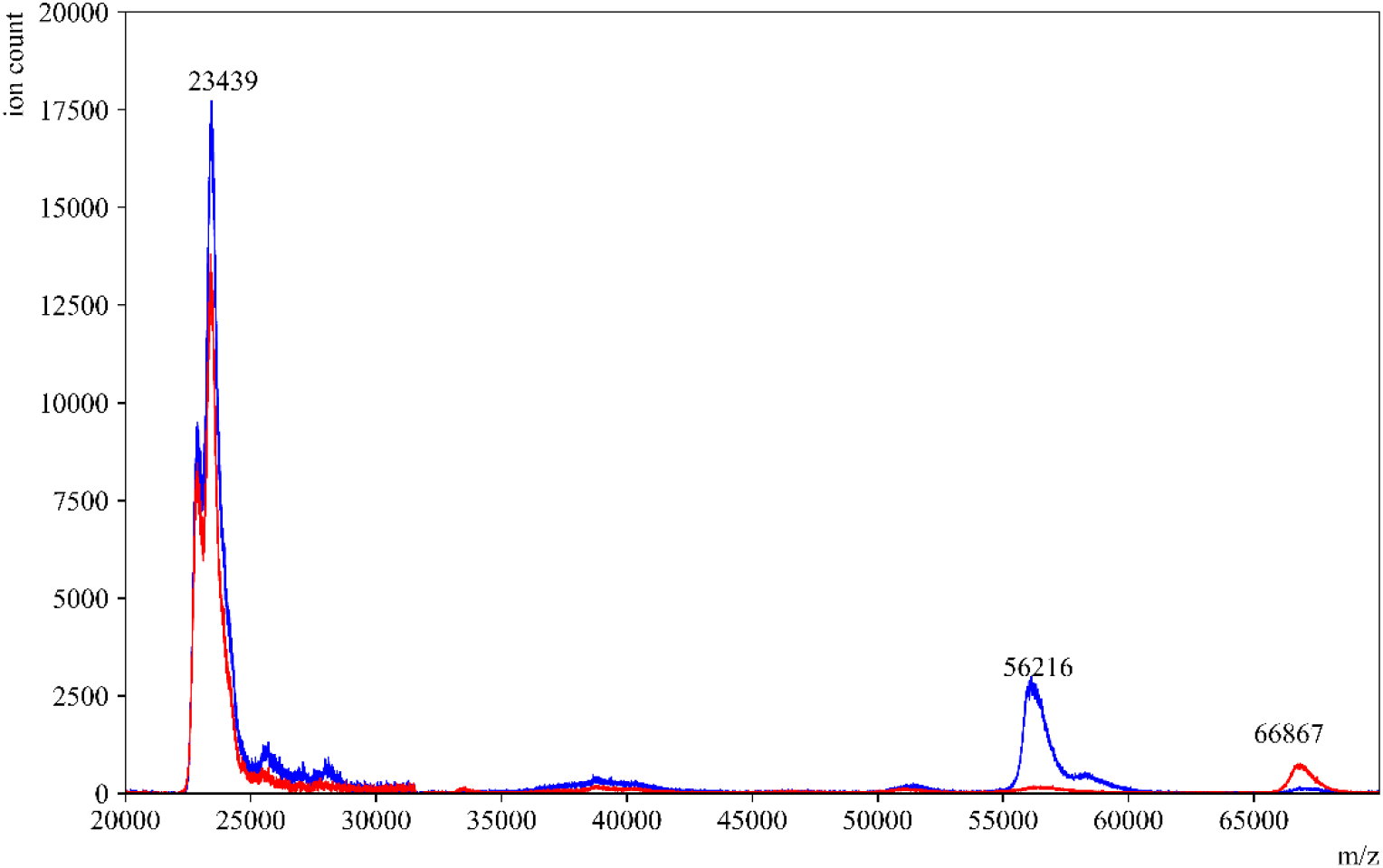
COVID-19 negative control saliva before and after amylase depletion. MALDI-ToF mass spectra of pre-COVID-19 saliva in (—blue) and an overlay of the same sample after passing through the amylase depleting device(—red). The peak at 56,216 m/z which corresponds to the presence of amylase, was significantly reduced in the sample’s eluent. Interestingly, the peak at 66,867 m/z was significantly enhanced in the eluent.

In order to further visualize the effects of passing gargle samples through the amylase depleting device, we performed an SDS-PAGE analysis. Figure 6 shows the positions of the bands for protein standards HSA, amylase and IgA. For IgA, two bands at ∼55 kDa and ∼25 kDa represent heavy and light chains respectively (Lane 4). Comparing the positions of these standard proteins, it can be distinctly observed that the band between 50-60 kDa in the gargle sample (Lane 5) decreased in intensity after passing through the amylase depleting device as seen in Lane 6. Furthermore, almost all the other bands were still observed in Lane 6. This coincides well with the corresponding MALDI-ToF spectra of the negative control, wherein the peak between 55,500–59,000 m/z significantly decreased in intensity upon passing the gargle samples through the amylase depleting device (Figure 5). As previously established, we observed that there is selective binding on the starch bed only w.r.t amylase, whereas all the other proteins showed a negligible loss in signal intensity on MALDI-ToF. Hence, we can conclude that the band getting depleted between 50-60 kDa in the SDS PAGE is corresponding to amylase. In Lane 6, we could still observe the light chains of immunoglobulins along with various other bands indicating that the rest of the proteome is conserved. Certainly, there is some protein loss occurring overall in the gargle samples while using the amylase depleting device as we are including an additional step in our analysis. However, when analyzing the eluents on a sensitive instrument such as MALDI-ToF, we can still observe all the other proteins even after depleting amylase.

**Figure 6:**
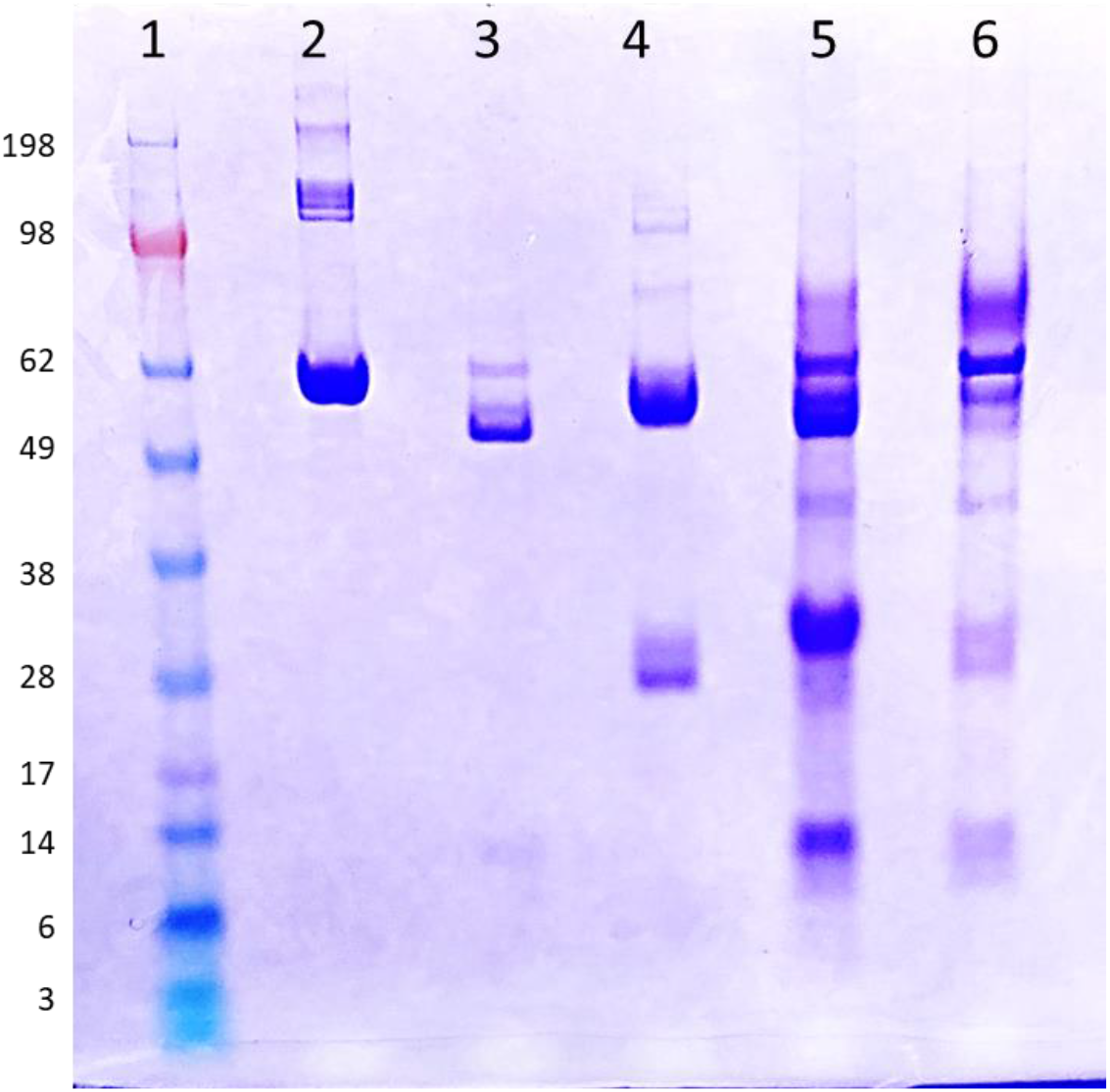
SDS-PAGE analysis of high-abundance proteins in human saliva and an example gargle sample stained with Coomassie-Brilliant-Blue under reducing conditions. Lane 1: Standard protein ladder. Lane 2: HSA (2.5 μg), Lane 3: salivary amylase (14 μg), Lane 4: IgA (3.5 μg). Lane 5: gargle sample before passing through amylase depleting device Lane 6: same gargle sample after passing through the device (eluent).

### The starch bed depletes amylase from saliva and unmasks other peaks

In gargles from subjects, we observed a substantial signal drop between 55,500–59,000 m/z in the MALDI-ToF profiles of COVID-19 positive samples after passage through the set-up (Figure 7). This suggests that the ions contributing to this peak are of amylase and are reduced in number upon employing the amylase depleting device. Moreover, the peak around 66,000 m/z (along with several other peaks) was remarkably enhanced in these eluents, supporting the hypothesis that amylase depletion unmasked signals from multiple proteins in saliva.

**Figure 7:**
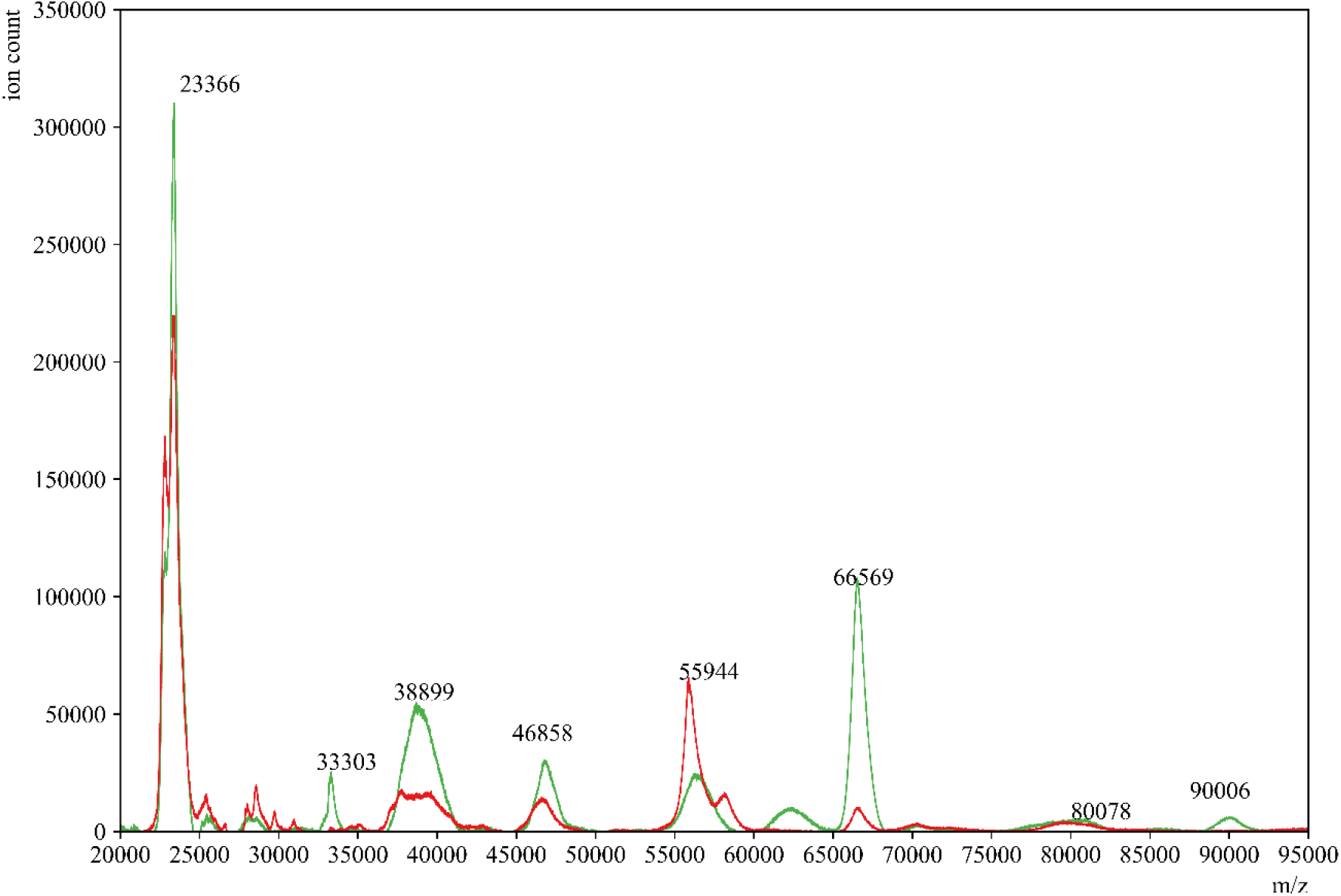
COVID-19 positive gargle sample before and after amylase depletion. MALDI-ToF mass spectra of a representative gargle sample from a COVID-19 positive donor (—red) and an overlay of the same sample passed through the amylase depleting device (—green).

Next, we compared the eluents of COVID-19 positive and COVID-19 negative gargle samples (Figure 8). As predicted, COVID-19 positive profiles had numerous and higher intensity peaks compared to COVID-19 negative profiles, even after depleting amylase. Specifically, the presumed viral protein peaks at the previously established ranges of 66,400–68,100 and 78,600–80,500 m/z were still significantly higher in COVID-19 positive compared to COVID-19 negative gargles suggesting that amylase depletion did not affect the ability to detect viral proteins in the spectra. SDS-PAGE analysis was also performed to compare the clinically confirmed COVID-19 positives and negative samples (Figure S1). The eluents of both of these samples (Lane 3 and Lane 5) showed a reduction in band intensity at around 60kDa which is close to the theoretical weight of salivary amylase. Moreover, the rest of the proteome was protected and could be visible by CBB staining method.

**Figure 8:**
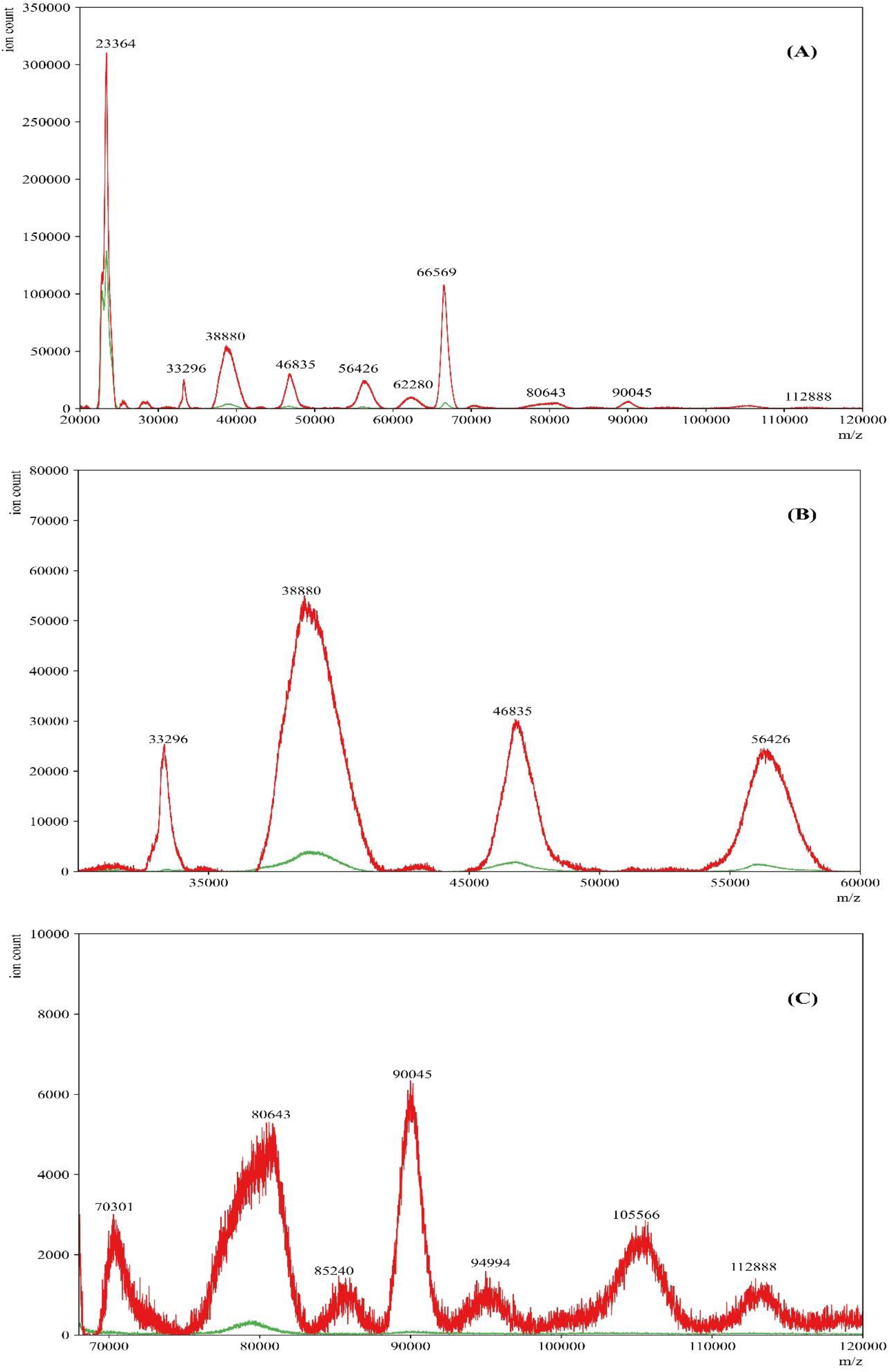
COVID-19 positive and negative gargle sample after amylase depletion. Representative MALDI-ToF mass spectra of gargle samples from a COVID-19 positive donor eluent (—red) and COVID-19 negative donor eluent (—green). Panel A is the full-range mass spectra of the samples. Panels B and C show specific ranges where differences in mass spectra are observed between COVID-19 positive eluent and COVID-19 negative eluent samples after amylase depletion.

### Sensitivity and specificity of SARS-COV-2 detection after amylase depletion

The sample files were normalized by dividing the signal intensities (from 2,000-200,00 m/z) by the peak intensity of the (M+H)^+^ peak of apomyoglobin which was used to calibrate the instrument daily. Next, the areas of the peaks in the 11,140– 11,160 m/z (quality control peak present in saliva irrespective of COVID-19 status) and 78,600-80,500 m/z (S1 spike protein of SARS-CoV-2) ranges were calculated by leveraging the composite Simpson’s rule for each spectral range as described in our previous study.[38] Simpson’s rule is a numerical method for approximating the definite integral of a function which is based on approximating the AUC using quadratic polynomials. The formula for Simpson’s rule involves dividing the interval of integration into an even number of subintervals and evaluating the function at the endpoints and midpoints of these subintervals. The approximate value of the integral is then given by a weighted sum of these function values.[42] ROC curve analysis was performed to determine the appropriate cut-off to call a sample COVID-19 positive vs negative. The disease prevalence for generating the ROC plot was assumed to be 10% based on the CDC’s COVID-19 data tracker. As expected, peaks in the 11,140–11,160 m/z range did not discriminate between COVID-19 negative and positive samples (AUC 0.77, threshold of 4.44).

Peaks in the range of 78,600-80,500 m/z that represent the S1 spike protein of SARS-CoV-2 showed good discrimination between COVID-19 positive and negative specimens (AUC 0.893, threshold of 10.14) after amylase depletion. Using a threshold area of 10.14, the sensitivity and specificity of COVID-19 detection was 85.71% and 100% respectively (Figure 9).

**Figure 9:**
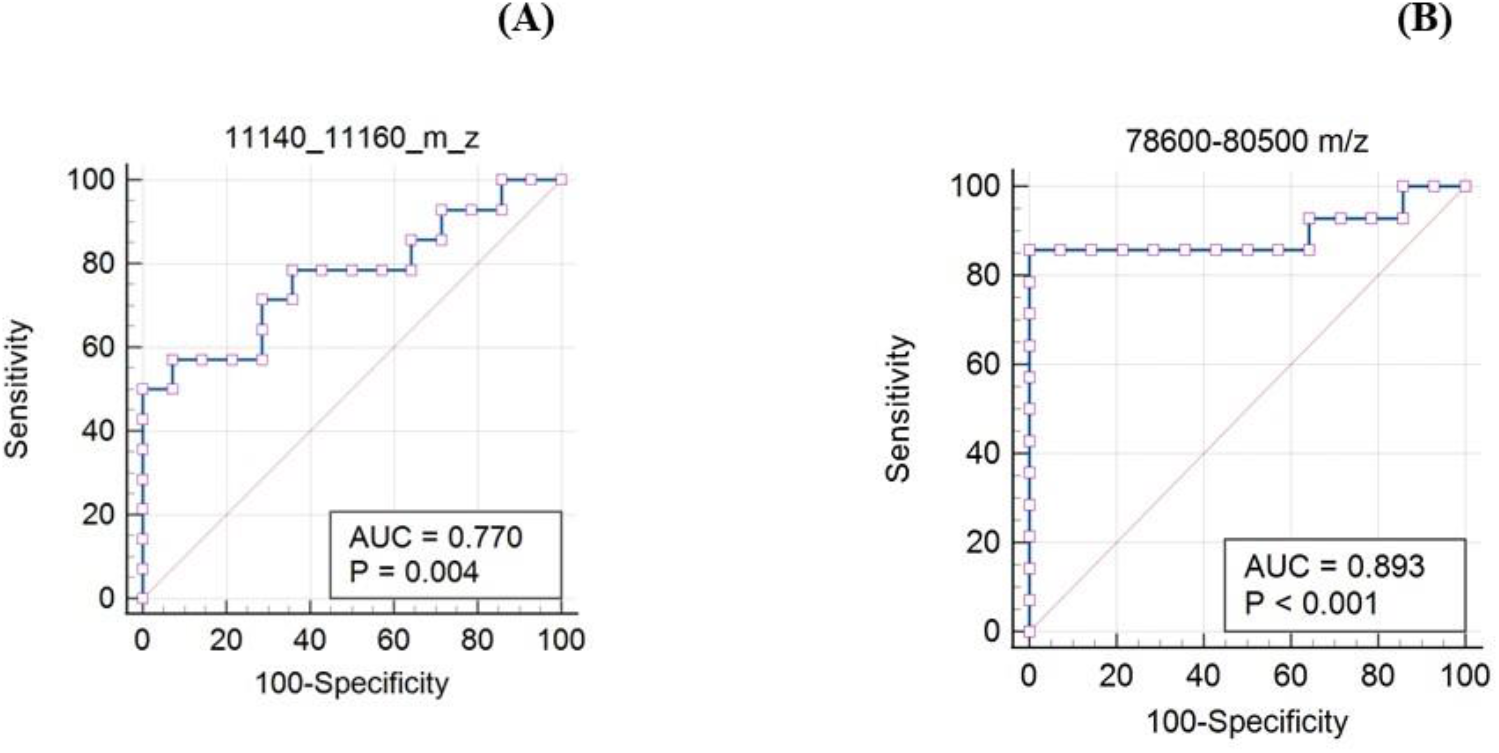
ROC analysis of the biomarker ranges 11,140-11,160 m/z and 78,600-80,500 m/z after amylase depletion. In the plot, each point represents a MALDI-ToF specificity/sensitivity value at a cutoff AUC for each potential biomarker range, with the highest specificity/sensitivity and discriminating power having values near the top left of each plot.

The sensitivity of SARS-CoV-2 detection after amylase depletion was somewhat lower than expected when compared to RT-PCR. It is quite possible that some of the positive samples we selected had low viral loads. IDPH reports positive cases as “detected” but does not report a Ct value to assess the level of the viral load. Further studies with a larger number of samples and comparison with Ct values of positive samples are necessary to estimate the realistic sensitivity of MALDI-ToF method to detect SARS-COV-2.

To assess how the amylase depletion process affected the sensitivity and specificity of our method, we analyzed the AUC values of the two peaks (11,140-11,160 m/z and 78,600-80,500 m/z) for the same set of samples before amylase depletion (Figure 10). There certainly was an improvement in the detection of COVID-19 after depleting amylase, as the specificity *before* amylase depletion was 57.12% whereas the sensitivity was still the same (85.71%). This again directs us towards the hypothesis that the signal intensity for the biomarkers of the disease (in this case 78,600-80,500 m/z) were being suppressed due to the abundance of amylase in the gargle samples.

**Figure 10:**
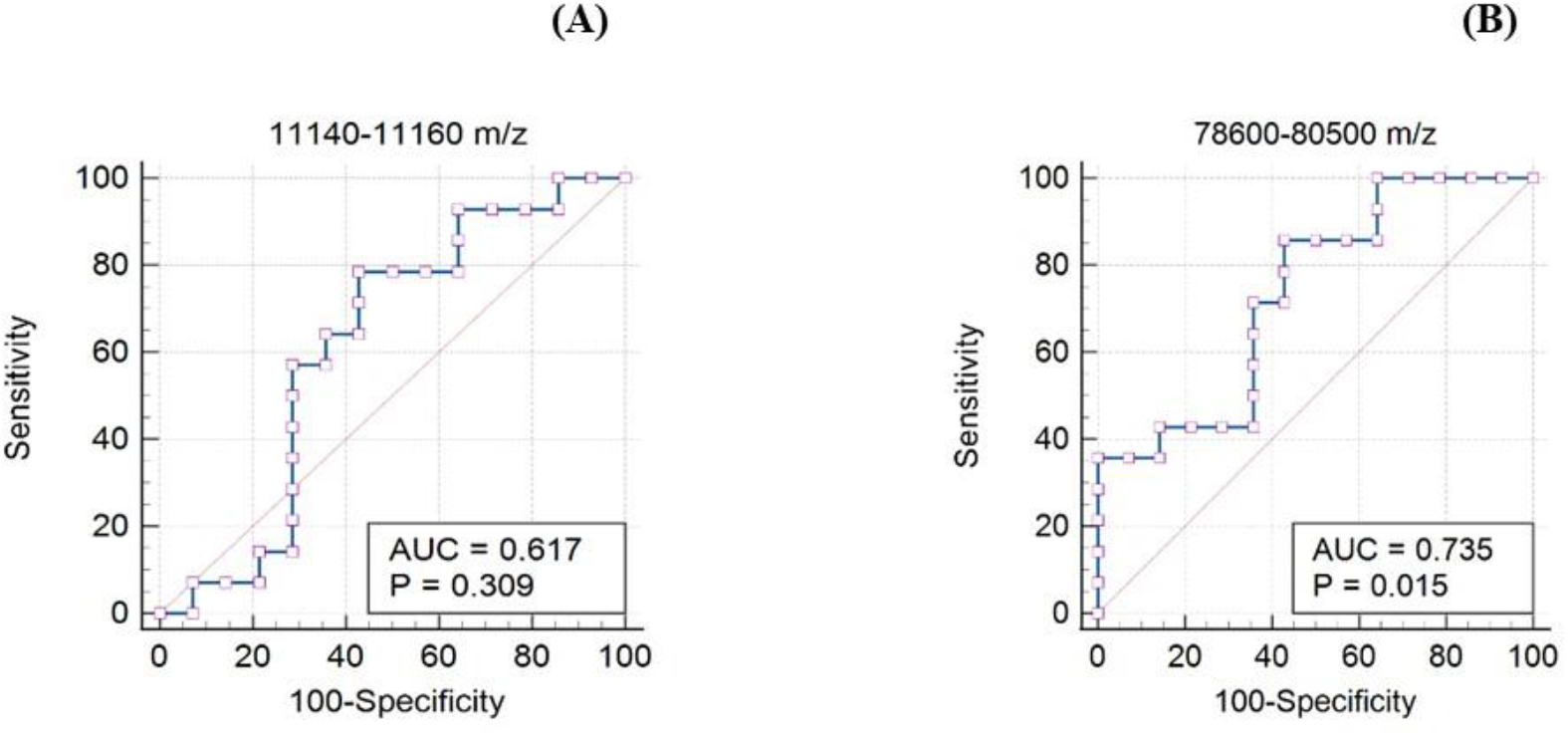
ROC analysis of the biomarker ranges 11,140-11,160 m/z and 78,600-80,500 m/z before amylase depletion. In the plot, each point represents a MALDI-ToF specificity/sensitivity value at a cutoff AUC for each potential biomarker range, with the highest specificity/sensitivity and discriminating power having values near the top left of each plot.

## Discussion

A key issue in proteomic analysis of any specimen by MALDI-ToF MS is the presence of high-abundance proteins which, in terms of assay development, can significantly mask the detection of other low-abundance proteins. This poses a hurdle to biomarker discovery as these low-abundance proteins might be potential candidates for use in the diagnosis of diseases. While there are various studies to deplete high-abundance proteins from blood, there exists limited data concerning saliva samples. One of the high-abundance proteins in saliva is amylase. Herein, we adopted a simple, cost-effective yet efficient method to deplete salivary amylase in COVID-19 gargle samples before MALDI-ToF MS analysis.

Deutsch et al. designed an amylase depleting device that included a 0.45 μm paper filter at the tip of the syringe.[34] In our initial efforts in preparing and utilizing this apparatus, we experienced difficulty while removing the plunger between starch activation and sample loading. Therefore, we replaced the paper filter with a 0.45 μm syringe filter (Figure 1), minimizing disturbances to the potato starch bed while releasing the plunger. We also passed increased amounts of water through the device before loading samples in order to remove water-soluble residues present within the potato starch that might contribute to the ion suppression of proteins or interfere with amylase adsorption (Figure 11). The presence of starch can be confirmed by the iodine reagent, which turns the sample deep blue in the presence of starch. It can be distinctly observed in Panel A of Figure 11 that starch was no longer present after passing 20 mL of water. This was also verified by the UV-Vis spectrometer shown in Figure 11 Panel B. Hence, 20 mL of water was passed through the amylase depleting device before passing any gargle sample or standards/controls as it resulted in a significant decrease in water-soluble residues present in starch. These additional washing steps were crucial to prevent any ion suppression of proteins in the MALDI-ToF analysis due to the presence of water-soluble starch residues in the samples. Research concerning the types of starch used and the influence of its structure on the interaction with amylase is poorly understood. Furthermore, it is stated that the Michaelis-Menten model cannot be applied to in-vitro studies of amylase action because of uncertainties in the structure of its substrate (starch), the possible presence of inhibitors and high enzymatic activity.[43],[44]

**Figure 11:**
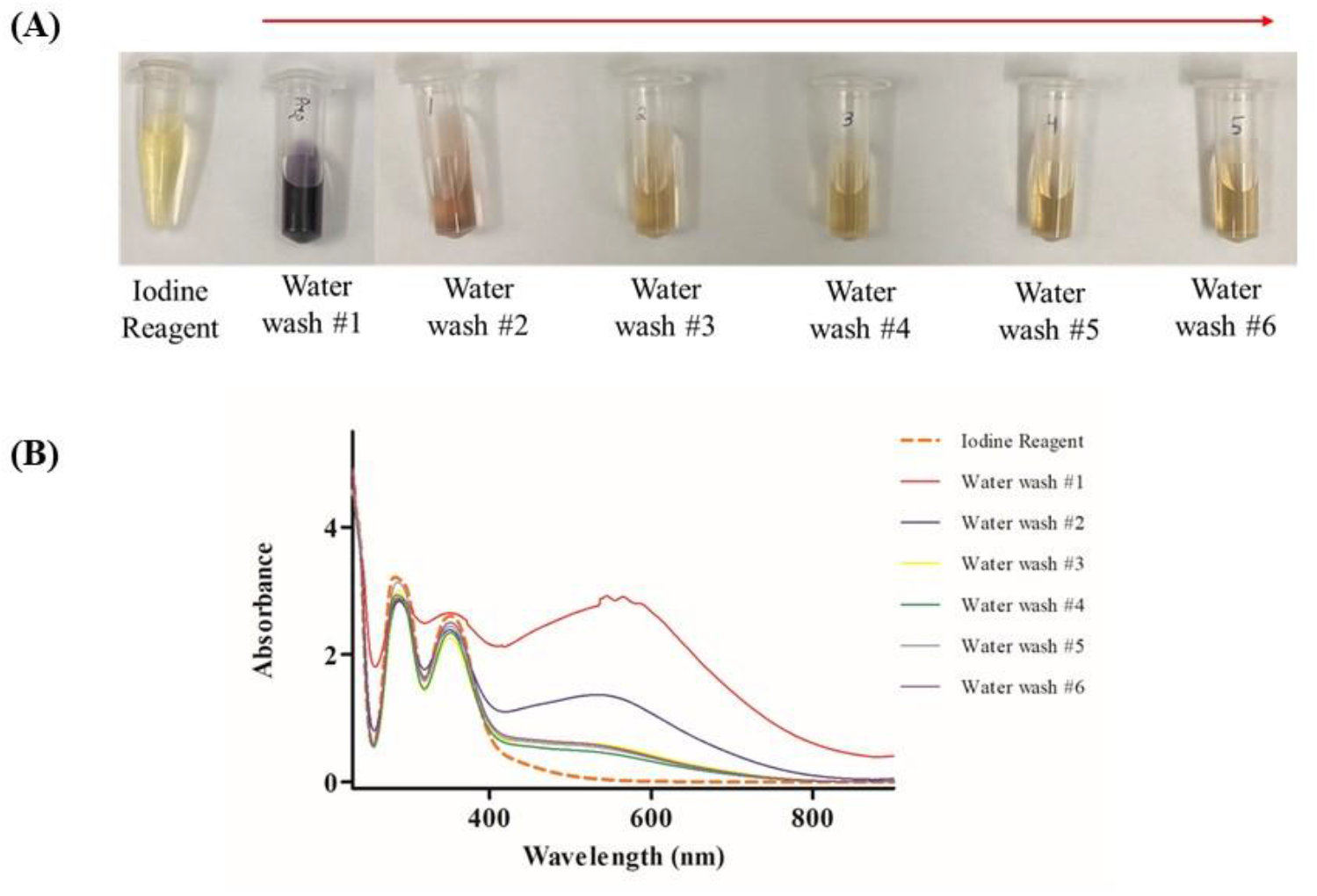
Iodine test to determine the presence of starch after multiple water washes. The gradual color change in Panel A and the overall decrease in the absorbance in the UV-Vis spectra in Panel B confirmed the reduction in water-soluble starch residues present in starch.

In previous studies, we used LBSD-X (MAPSciences Bedford, UK) buffer to lyse viral particles in saliva. However, supply chain issues have hampered the availability of this buffer. This motivated us to utilize a previously reported buffer for viral reconstitution.[39] We optimized the buffer by substituting 2 mM dithiothreitol with 50 mM TCEP and by substituting sonication followed by 2-hour incubation with a simple vortex and 15-minute incubation at room temperature. The buffer showed promising results as we observed peaks within the ranges established previously (11,140–11,160 m/z; 23,550– 23,800 m/z; 27,900–29,400 m/z; 55,500–59,000 m/z; 66,400–68,100 m/z; 78,600–80,500 m/z; and 111,500–115,500 m/z; Figure 12). Furthermore, and perhaps more importantly, we could still observe differences between COVID-19 positive and negative gargle samples (Figure 8).

**Figure 12:**
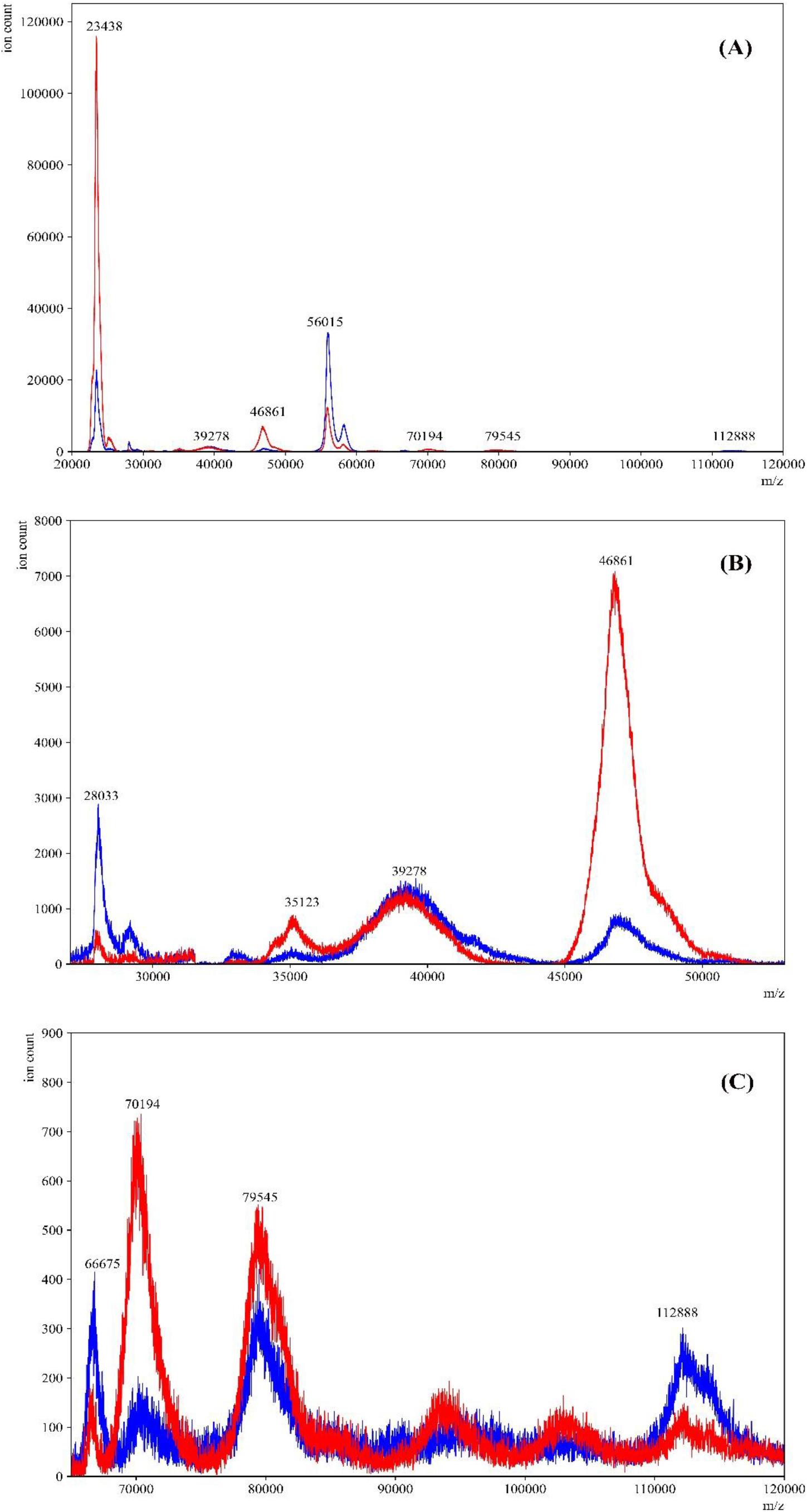
Comparison between the LBSD-X buffer and the newly optimized viral disruption buffer. MALDI-ToF mass spectra of a COVID-19 positive gargle treated with LBSD-X buffer (—red) and an overlay of the same sample treated with the new optimized buffer (—blue). Panel A is the full-range mass spectra of the same positive sample treated with both buffers. The optimized buffer showed peaks in the ranges that were established in our previous study. Panels B and C are specific ranges in mass spectra where the peaks obtained by both the buffers closely overlap with each other.

After comparing peak areas between the eluents of COVID-19 positive and negative gargle samples, we found that the samples could be discriminated by the 78,600–80,500 m/z range which corresponds to the theoretical m/z of the S1 fragment of the spike protein of SARS-CoV-2. Additionally, we performed an ROC curve analysis to establish a cut-off for the peak area to assess diagnostic utility. A threshold of >10.14 units for 78,600–80,500 m/z range resulted in a specificity and sensitivity of 100% and 85.17% respectively for COVID-19 positivity.

Even though five biomarkers were established in our previous study, we only chose to analyze samples based on the 78,600– 80,500 m/z range. The biomarkers in the range of 55,500–59,000 m/z and 27,900–29,400 m/z are the singly and double charged peaks of IgA Heavy chain+Amylase. These peaks seem to be depleted after using the amylase depleting device as the starch bed shows affinity toward the amylase. Moreover, as shown in Figures 2 and 4, when the standard IgA and amylase were passed individually on the amylase depleting device, we could see the selective binding only towards amylase and minimal difference in the intensities for IgA. Hence, when considering a gargle sample, it is important to note the presence of two proteins in the ranges of 55,500–59,000 m/z and 27,900–29,400 m/z amongst which only one protein (amylase) is getting depleted. Overall, there certainly is some protein loss while using this device. But irrespective of this loss, we could see the effects of amylase depletion on the intensities of other proteins. The S2 fragment of the Spike protein of SARS-CoV-2 (66,400–68,100 m/z) was not considered in this study because it overlaps with the albumin (∼66,348 kDa) peak present in the host saliva. Nevertheless, some viral proteins are still retained after amylase depletion and have the potential to be used as biomarkers of disease processes in saliva.

Several eluents of gargle samples from amylase-depleted samples showed the presence of additional peaks apart from our established biomarker ranges that were suppressed in non-depleted samples due to the ions from amylase. Further studies are needed to characterize these peaks and determine their utility in diseased and normal conditions.

Finally, previous potato starch column studies were successful in depleting salivary amylase but were not applied towards clinical applications. For the first time, we report a potential application of this method for the diagnosis of COVID-19. Because amylase depletion unmasks peaks that were otherwise not visible, the quick and cost-efficient method can be employed to discover novel biomarkers for diseases by MALDI-ToF analysis of saliva. Our proof-of-concept study shows that other high abundance proteins can be potentially depleted in a similar manner to improve the diagnostic utility of saliva. Overall, this type of sample processing can result in improved sensitivities of protein analytes present in low abundance.

## Conclusion

An inexpensive device to deplete salivary amylase from human gargle samples was employed to improve the protein profiles of COVID-19 positive and negative samples in saliva. The starch bed selectively captured salivary amylase whereas the rest of the proteome remained largely unaffected when passed through the amylase depleting device. Furthermore, this method resulted in enhanced MALDI-ToF signals of various other peaks which were suppressed by the presence of amylase. Overall, a sensitivity of 85% and specificity of 100% were achieved for a total of 28 gargle samples (14 COVID-19 positives and COVID-19 negatives samples) by ROC curve analysis. Biomarkers that are usually present in low abundance could plausibly be unmasked and detected by utilizing such a technique.

## Data Availability

All de-identified data produced in the present study are available upon reasonable request to the authors.

## Abbreviations

ACE-2: angiotensin-converting enzyme 2
AUC: area under the curve
COVID-19: coronavirus disease 2019
DTT: dithiothreitol
HSA: human serum albumin
IDPH: Illinois Department of Public Health
IgA: immunoglobulin A
LC-MS: liquid chromatography-mass spectrometry
MALDI-ToF MS: matrix-assisted laser desorption/ionization-time of flight mass spectrometry
NP: nasopharyngeal
RNA: ribonucleic acid
ROC: receiver operating characteristic
RT-qPCR: reverse transcriptase quantitative polymerase chain reaction
S protein: spike protein
SARS-CoV-2: severe acute respiratory syndrome coronavirus 2
SDS-PAGE: sodium dodecyl sulfate polyacrylamide gel electrophoresis

## Declaration of Competing Interest

The authors declare they have no known competing financial interests or personal relationships that could affect the work described in this article.

## Acknowledgements

This work was supported by Northern Illinois University’s Molecular Analysis Core which was established with support from Shimadzu Scientific Instruments. We would also like to thank Dr. Pratool Bharti (Intel Corporation, Oregon, USA) for helpful discussions on data analysis.

## IRB information

The protocol “Use of MALDI TOF mass spectrometry to analyze SARS CoV2 viral proteins” was approved on August 12, 2020, and revised on July 12, 2021, and August 11, 2022 by Northern Illinois University.

## Notes

### Competing Interest Statement

The authors have declared no competing interest.

### Funding Statement

This study did not receive any funding

### Author Declarations

Institutional Review Board and Institutional Biosafety Committee of Northern Illinois University (approved on August 12, 2020, and revised on July 12, 2021, and August 11, 2022)

## References

[1] K. E. Kaczor-Urbanowicz, C. Martin Carreras-Presas, K. Aro, M. Tu, F. Garcia-Godoy, and D. T. W. Wong, “Saliva diagnostics – Current views and directions,” Experimental Biology and Medicine, vol. 242, no. 5, pp. 459–472, Mar. 2017. doi: 10.1177/1535370216681550.

[2] K. T. B. Crosara, D. Zuanazzi, E. B. Moffa, Y. Xiao, M. A. D. A. M. MacHado, and W. L. Siqueira, “Revealing the Amylase Interactome in Whole Saliva Using Proteomic Approaches,” Biomed Research International, vol. 2018, Jan. 2018, doi: 10.1155/2018/6346954.

[3] G. Krief, O. Deutsch, B. Zaks, D. T. Wong, D. J. Aframian, and A. Palmon, “Comparison of diverse affinity based high-abundance protein depletion strategies for improved bio-marker discovery in oral fluids,” Journal of Proteomics, vol. 75, no. 13, pp. 4165–4175, Jul. 2012, doi: 10.1016/j.jprot.2012.05.012.

[4] C. Z. Zhang et al., “Saliva in the diagnosis of diseases,” International Journal of Oral Science, vol. 8, pp. 133–137, Sep. 2016. doi: 10.1038/ijos.2016.38.

[5] P. N. Floriano et al., “Use of saliva-based nano-biochip tests for acute myocardial infarction at the point of care: A feasibility study,” Clinical Chemistry, vol. 55, no. 8, pp. 1530–1538, Aug. 2009, doi: 10.1373/clinchem.2008.117713.

[6] S. Bencharit et al., “Salivary proteins associated with hyperglycemia in diabetes: A proteomic analysis,” Molecular Biosystems, vol. 9, no. 11, pp. 2785–2797, Sept. 2013, doi: 10.1039/c3mb70196d.

[7] Q. Wang, P. Gao, X. Wang, and Y. Duan, “The early diagnosis and monitoring of squamous cell carcinoma via saliva metabolomics,” Scientific Reports, vol. 4, Oct. 2014, doi: 10.1038/srep06802.

[8] F. Wei et al., “Noninvasive saliva-based EGFR gene mutation detection in patients with lung cancer,” American Journal of Respiratory and Critical Care Medicine, vol. 190, no. 10, pp. 1117–1126, Nov. 2014, doi: 10.1164/rccm.201406-1003OC.

[9] K. Gao et al., “Systemic disease-induced salivary biomarker profiles in mouse models of melanoma and non-small cell lung cancer,” PLoS One, vol. 4, no. 6, Jun. 2009, doi: 10.1371/journal.pone.0005875.

[10] N. L. Rhodus, B. Cheng, S. Myers, L. Miller, V. Ho, and F. Ondrey, “The feasibility of monitoring NF-κB associated cytokines: TNF-α, IL-1α, IL-6, and IL-8 in whole saliva for the malignant transformation of oral lichen planus,” Molecular Carcinogenesis, vol. 44, no. 2, pp. 77–82, Mar. 2005, doi: 10.1002/mc.20113.

[11] J. J. Ceron et al., “Use of saliva for diagnosis and monitoring the SARS-CoV-2: A general perspective,” Journal of Clinical Medicine, vol. 9, no. 5, May 2020. doi: 10.3390/jcm9051491.

[12] T. Sri Santosh, R. Parmar, H. Anand, K. Srikanth, and M. Saritha, “A Review of Salivary Diagnostics and Its Potential Implication in Detection of Covid-19,” Cureus, Apr. 2020, doi: 10.7759/cureus.7708.

[13] K. Elżbieta Kaczor-Urbanowicz, “Salivary Diagnostics,” in Salivary Glands - New Approaches in Diagnostics and Treatment, IntechOpen, 2019. doi: 10.5772/intechopen.73372.

[14] J.-L. Capelo-Martinez, “Emerging Sample Treatments in Proteomics”, Springer, doi:10.1007/978-3-030-12298-0.

[15] N. M. O’Brien-Simpson, K. Burgess, G. C. Brammar, I. B. Darby, and E. C. Reynolds, “ Development and evaluation of a saliva-based chair-side diagnostic for the detection of Porphyromonas gingivalis, ” Journal of Oral Microbiology, vol. 7, no. 1, p. 29129, Jan. 2015, doi: 10.3402/jom.v7.29129.

[16] J. L. Ebersole, R. Nagarajan, D. Akers, and C. S. Miller, “Targeted salivary biomarkers for discrimination of periodontal health and disease(s),” Frontiers Cellular and Infection Microbiology, vol. 5, no. Aug, 2015, doi: 10.3389/fcimb.2015.00062.

[17] C. F. Streckfus, L. R. Bigler, and M. Zwick, “The use of surface-enhanced laser desorption/ionization time-of-flight mass spectrometry to detect putative breast cancer markers in saliva: a feasibility study,” Journal of Oral Pathol Med, vol.35, pp.292–300, 2006.

[18] Y. J. Jou et al., “Salivary zinc finger protein 510 peptide as a novel biomarker for detection of oral squamous cell carcinoma in early stages,” Clinica Chimica Acta, vol. 412, no. 15–16, pp. 1357–1365, Jul. 2011, doi: 10.1016/j.cca.2011.04.004.

[19] N. Delaleu, P. Mydel, I. Kwee, J. G. Brun, M. v. Jonsson, and R. Jonsson, “High fidelity between saliva proteomics and the biologic state of salivary glands defines biomarker signatures for primary Sjögren’s syndrome,” Arthritis and Rheumatology, vol. 67, no. 4, pp. 1084–1095, Apr. 2015, doi: 10.1002/art.39015.

[20] M. Castagnola et al., “Hypo-phosphorylation of salivary peptidome as a clue to the molecular pathogenesis of autism spectrum disorders,” Journal of Proteome Research, vol. 7, no. 12, pp. 5327–5332, Dec. 2008, doi: 10.1021/pr8004088.

[21] S. V. Khadse, G. Bajaj, P. Vibhakar, P. Nainani, R. Ahuja, and G. Deep, “Evaluation of specificity and sensitivity of oral fluid for diagnosis of hepatitis B,” Journal of Clinical and Diagnostic Research, vol. 10, no. 1, pp. BC12–BC14, Jan. 2016, doi: 10.7860/JCDR/2016/17319.7107.

[22] S. P. Sweet, A. N. Denbury, and S. J. Challacombe, “Salivary calprotectin levels are raised in patients with oral candidiasis or Sjogren’s syndrome but decreased by HIV infection,” Oral Microbiology and Immunology, vol. 16, pp. 119–123, 2001.

[23] A. C. Andries et al., “Value of Routine Dengue Diagnostic Tests in Urine and Saliva Specimens,” PLoS Neglected Tropical Diseases, vol. 9, no. 9, Sep. 2015, doi: 10.1371/journal.pntd.0004100.

[24] K. R. Katsani and D. Sakellari, “Saliva proteomics updates in biomedicine,” Journal of Biological Research (Thessaloniki), vol. 26, no. 1, Jan. 30, 2019. doi: 10.1186/s40709-019-0109-7.

[25] I. Duś-Ilnicka, E. Krala, P. Cholewińska, and M. Radwan-Oczko, “The use of saliva as a biosample in the light of COVID-19,” Diagnostics, vol. 11, no. 10, Sept. 2021. doi: 10.3390/diagnostics11101769.

[26] D. C. Caixeta et al., “One-Year Update on Salivary Diagnostic of COVID-19,” Frontiers in Public Health, vol. 9, May 2021. doi: 10.3389/fpubh.2021.589564.

[27] L. M. Czumbel et al., “Saliva as a candidate for COVID-19 diagnostic testing: A meta-analysis,” Frontiers in Medicine, vol. 7, Aug. 2020. doi: 10.3389/fmed.2020.00465.

[28] E. Borghi, V. Massa, G. Zuccotti, and A. L. Wyllie, “Testing Saliva to Reveal the Submerged Cases of the COVID-19 Iceberg,” Frontiers Microbiology, vol. 12, Jul. 2021, doi: 10.3389/fmicb.2021.721635.

[29] G. Krief, O. Deutsch, S. Gariba, B. Zaks, D. J. Aframian, and A. Palmon, “Improved visualization of low abundance oral fluid proteins after triple depletion of alpha amylase, albumin and IgG,” Oral Diseases, vol. 17, no. 1, pp. 45–52, Jan. 2011, doi: 10.1111/j.1601-0825.2010.01700.x.

[30] T. Do, R. Guran, V. Adam, and O. Zitka, “Use of MALDI-TOF mass spectrometry for virus identification: a review,” Analyst, vol. 147, no. 14, pp. 3131–3154, Jun. 2022. doi: 10.1039/d2an00431c.

[31] M. D. Contreras-Aguilar, S. v. Mateo, F. Tecles, C. Hirtz, D. Escribano, and J. J. Cerón, “Changes occurring on the activity of salivary alpha-amylase proteoforms in two naturalistic situations using a spectrophotometric assay,” Biology (Basel), vol. 10, no. 3, Mar. 2021, doi: 10.3390/biology10030227.

[32] C. Hirtz et al., “MS characterization of multiple forms of alpha-amylase in human saliva,” Proteomics, vol. 5, no. 17, pp. 4597–4607, Mar. 2005, doi: 10.1002/pmic.200401316.

[33] C. Peyrot des Gachons and P. A. S. Breslin, “Salivary Amylase: Digestion and Metabolic Syndrome,” Current Diabetes Reports, vol. 16, no. 10, Oct. 2016. doi: 10.1007/s11892-016-0794-7.

[34] O. Deutsch, Y. Fleissig, B. Zaks, G. Krief, D. J. Aframian, and A. Palmon, “An approach to remove alpha amylase for proteomic analysis of low abundance biomarkers in human saliva,” Electrophoresis, vol. 29, no. 20, pp. 4150–4157, May 2008, doi: 10.1002/elps.200800207.

[35] S. Bano, S. A. U. Qader, A. Aman, M. N. Syed, and A. Azhar, “Purification and characterization of novel α-amylase from bacillus subtilis KIBGE HAS,” American Association of Pharmaceutical Scientists, vol. 12, no. 1, pp. 255–261, Jan. 2011, doi: 10.1208/s12249-011-9586-1.

[36] S. Hyung, G. Karima, K. Shin, K. S. Kim, and J. W. Hong, “A Simple Paper-Based α-Amylase Separating System for Potential Application in Biological Sciences,” Biochip Journal, vol. 15, no. 3, pp. 252–259, Feb. 2020, doi: 10.1007/s13206-021-00022-3.

[37] H. Xiao and D. T. W. Wong, “Method development for proteome stabilization in human saliva,” Analytica Chimica Acta, vol. 722, pp. 63–69, Feb. 2012, doi: 10.1016/j.aca.2012.02.017.

[38] P. Chivte et al., “MALDI-ToF protein profiling as a potential rapid diagnostic platform for COVID-19,” Journal of Mass Spectrometry and Advances in the Clinical Lab, vol. 21, pp. 31–41, Aug. 2021, doi: 10.1016/j.jmsacl.2021.09.001.

[39] N. L. Dollman, J. H. Griffin, and K. M. Downard, “Detection, Mapping, and Proteotyping of SARS-CoV-2 Coronavirus with High Resolution Mass Spectrometry,” ACS Infectious Diseases, vol. 6, no. 12, pp. 3269–3276, Sept. 2020, doi: 10.1021/acsinfecdis.0c00664.

[40] A. B. Schwahn, J. W. H. Wong, and K. M. Downard, “Signature peptides of influenza nucleoprotein for the typing and subtyping of the virus by high resolution mass spectrometry,” Analyst, vol. 134, no. 11, pp. 2253–2261, Aug. 2009, doi: 10.1039/b912234f.

[41] A. B. Schwahn, J. W. H. Wong, and K. M. Downard, “Typing of human and animal strains of influenza virus with conserved signature peptides of matrix M1 protein by high resolution mass spectrometry,” Journal of Virological Methods, vol. 165, no. 2, pp. 178–185, Jan. 2010, doi: 10.1016/j.jviromet.2010.01.015.

[42] R. J. T. B. Murray, Area under a Curve: Trapezoidal and Simpson’s Rules. 1987. doi: https://doi.org/10.1007/978-1-4612-4974-0_26.

[43] F. J. Warren, P. G. Royall, S. Gaisford, P. J. Butterworth, and P. R. Ellis, “Binding interactions of α-amylase with starch granules: The influence of supramolecular structure and surface area,” Carbohydrate Polymers, vol. 86, no. 2, pp. 1038–1047, May 2011, doi: 10.1016/j.carbpol.2011.05.062.

[44] P. J. Butterworth, F. J. Warren, and P. R. Ellis, “Human α-amylase and starch digestion: An interesting marriage,” Starch/Staerke, vol. 63, no. 7. pp. 395–405, Jan. 2011. doi: 10.1002/star.201000150.

