## Supplementary information for "AMYLASE DEPLETION SIGNIFICANTLY IMPROVES THE MALDI-TOF PROFILE OF SALIVA – APPLICATION TO COVID-19 DIAGNOSTICS"

#co-first authors

*corresponding author

**Supplementary information:**

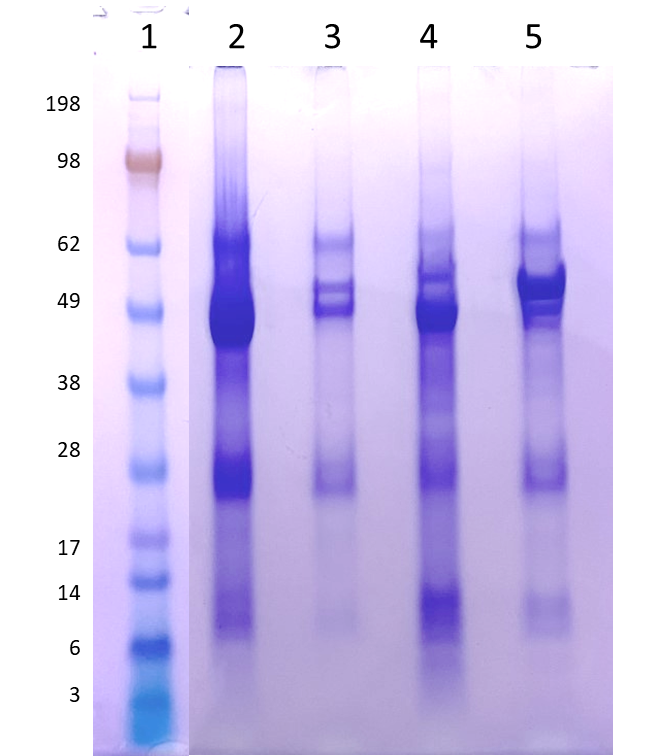

***Figure S1: SDS-PAGE analysis of COVID-19 positive and COVID-19 negative sample and the respective eluents.***

*Lane 1:Standard protein ladder, Lane 2: COVID-19 positive sample before passing through the amylase depleting device while Lane 3 represents its eluent. Lane 4 represents the COVID-19 negative sample whereas Lane 5 represents its eluent.*
